# All cause mortality, re-amputation and readmission rates following below knee amputation in 24,711 diabetic patients; a 25-year England population study (1998 to 2023)

**DOI:** 10.1101/2025.03.10.25323685

**Authors:** Conor Hennessy, Simon Abram, Rick Brown, Constantinos Loizou, Robert Sharp, Adrian Kendal

## Abstract

**Objectives:** To study changes in all cause mortality of patients with diabetes following below knee amputation (BKA) in England from 1998 to 2023; identifying at risk patients. To equip patients and healthcare providers with accurate estimates of serious adverse events including 90-day complications, reoperation, readmission and further lower limb amputation rates.

**Design:** National population based cohort study.

**Setting:** Hospital Episode Statistics database for NHS England linked to ONS mortality records.

**Participants:** 24,711 patients with diabetes who underwent a below knee amputation in England between April 1998 and April 2023.

**Main outcome measures:** Primary outcomes were all cause mortality rate and amputation free survival calculated with Kaplan-Meier curve analyses. Multivariate logistic regression was used to stratify patient variables associated with mortality and/or re-amputation rate. Secondary outcomes included causes of death, re-amputation rates, temporal variation in post BKA mortality and rate of 90-day peri-operative complications and readmission rates.

**Results:** We identified 24,711 BKA on patients with diabetes in the 25-year period. The rate of BKA decreased from 2002 (5.9/100,000; 95% CI 5.6–6.1/100,000) to 2012 (4.4/100,000; 95% CI 4.2 – 4.5/100,000) and plateaued between 2012-2022 (4.3/100,000 in 2022; 95% CI 4.1 – 4.5/100,000). BKA rates were significantly higher in males (6.7/100,000; 95% CI 6.4 – 7/100,000) compared to females (2.0/100,000; 95% CI 1.8 – 2.1/100,000, P<0.05).

The mortality rates following BKA were 4.6% (30 days), 11.3% (90 days), 23.6% (1 year), and 58.2% (5 years). Only 13% of patients survived to 15 years. The commonest causes of death were chromic ischemic heart disease (16.3%), acute myocardial infarction (10.03%) and medical complications of diabetes (9.0%). Cox Proportional Hazard modelling found female patients (HR 1.08; 95% CI 1.05 – 1.12), increasing age (HR 1.79; 95% CI 1.73 – 1.86 in 60-79 year olds), and higher Charlson co-morbidity index (HR 2.60; 95% CI 1.74 – 3.88) were associated with significantly higher risk of mortality.

90-day post operative complications included reoperation (20.7%), myocardial infarction (3.2%), cerebrovascular accident (1.5%), and pulmonary embolus (0.59%). The ipsilateral re-amputation rate was 10.4% (n=2909), and the contralateral amputation rate was 11.4% (n=2304).

The average length of stay was 33 days (IQR 29, 1-152). 23.4% of patients were readmitted to hospital within 90 days.

**Conclusions:** This landmark 25-year England population study has revealed that BKA in patients with diabetes is still associated with high mortality rates, high rate of further amputation and high incidence of significant complications. Nearly a quarter of patients require readmission within 90 days. Severely co-morbid female patients over the age of 60 years have the highest mortality rate and represent an at risk group in need of national intervention.

## Introduction

Diabetes mellitus remains a major pandemic affecting 415 million people globally (9% of adults), of which 4 million are in the United Kingdom. 7% of patients with diabetes develop foot ulceration ^1 2^ and are 10 times more likely to require a lower limb amputation than those without diabetes ^3 4^. Major lower limb amputation, most commonly below knee amputation (BKA), remains a lifesaving treatment for severe diabetic foot disease.

The incidence of major amputation for diabetic foot disease decreased in England from 2003 to 2013^5^. Thereafter, the most recent United Kingdom National Diabetes Foot Care Audit (2014-21) found no further significant change in BKA rate following ‘first expert assessment’ of an ulcer^6^. This is despite reported improvements in ulcer healing rates following national investment in implementing NICE guidance for diabetic foot care^7^. What is not clear is if there has been an improvement in survival post BKA for diabetic foot disease.

Global data on BKA associated mortality is dominated by US Veterans population studies, comprising over 90% of published patient data^8^. 40% of patients in these studies do not have diabetes and almost all are male ^8–12^. Meta-analysis of a combined international dataset, including much smaller cohorts from 11 other countries, revealed a mean weighted mortality at 5 years of 65% and a wide range of 19% to 81%^12^. Historically, few long-term studies have focused solely on BKA in patients with diabetes, and of those that did, the highest number of patients was 1043^13^. Belgium reported the first national population study of BKA in patients with diabetes in 2023 and observed a 5-year mortality rate of 52% in 13,000 patients.

There is little clarity on the true mortality rate following amputation for diabetic foot disease in a United Kingdom national population^14^. The majority of UK larger studies and/or national population audits have focused on the outcomes of BKA for peripheral arterial disease (PAD)^15–17^. These include a recent study of geographical variation of major amputation rates for PAD in England that found diabetes was an independent risk factor for higher mortality^18^.

We lack a long-term England population based study of mortality post BKA for diabetic foot disease alone. It is also not clear if the mortality rates or causes of death post BKA in patients with diabetes have changed over the last 25 years.

This study uses Office of National Statistics (ONS) mortality linked to Hospital Episode Statistics data to examine temporal trends in all cause mortality following lower limb amputation in England for patients with diabetes. It seeks to identify those patients with diabetes who are most at risk of death, peri-operative complications, and rare but serious adverse events following BKA.

## Objectives

To study changes in all cause mortality of patients with diabetes following below knee amputation (BKA) in England from 1998 to 2023; identifying at risk patients. To equip patients and healthcare providers with accurate estimates of serious adverse events including 90-day complications, reoperation, readmission and further lower limb amputation rates.

## Methods

### Data source

We conducted a nationwide cohort study using data obtained from the Hospital Episodes Statistics (HES) database for England, combined with Office of National Statistics (ONS) mortality data^19^. The HES database holds information on all patients admitted to NHS hospitals in England. Each record in the database relates to one finished Consultant episode, describing the time an individual spends under the care of one NHS Consultant. Procedures performed in private hospitals are excluded. Submission of records to HES is mandated for accurate remuneration of NHS hospitals, and as such HES provides universal coverage of day case and inpatient surgical care. Any procedure performed is recorded using the Office of Population Censuses and Surveys Classification of Interventions and Procedures, version 4 (OPCS-4) codes. The World Health Organization International Classification of Diseases, 10th revision (ICD-10) codes are used to record diagnoses. The database includes detailed demographic data, co-morbidities, peri-operative complications and any further operative intervention. Hospital episode statistics data were then linked to mortality records from the Office for National Statistics, which provided information about the date and cause of death.

### Participants

Data was extracted on all patients over the age of 18 years with diabetes, who had undergone a below knee amputation (OPCS-4 listed in Supplementary Table 1) between April 2000 and April 2022. Patients were excluded if they had had a previous BKA, a more proximal amputation and/or if the BKA was for trauma, primary or secondary bone tumours.

### Data processing

The index event was defined as the first below knee amputation and data on all linkable episodes were extracted, including any prior hospital episode during the study period. Primary diagnoses and serious adverse events were identified based on specific ICD-10 codes. The Charlson comorbidity indices were calculated using the ICD-10 codes from HES records up to and including the index episode, using a previously validated algorithm^20^.

Duplicates, patients with incomplete records and/or chronologically inconsistent operative sequences were excluded. To avoid disclosure of personal identifiers, NHS Digital releases individual level data in a pseudonymised format with a restricted level of detail for demographic fields. Longitudinal follow-up of subsequent surgical interventions can be carried out by linking episodes through pseudonymised identifiers ^21^.

The HES data was linked to accurate records for cause and date of death from the Office for National Statistics.

### Statistical analysis

The primary outcome was the rate of all-cause mortality. Secondary outcome measures were causes of death, re-amputation rates, temporal variation in post BKA mortality and rate of 90-day peri-operative complications. Analysis was performed using R version 2.1 (R Foundation, Vienna, Austria) and STATA 18 (StataCorp, College station, Texas, USA). Descriptive statistics of patient demographics were analysed with Chi squared tests used for categorical variables and Wilcoxon Rank-Sum tests used for continuous variables. Mortality and amputation free survival was calculated with Kaplan-Meier curve analysis.

We used cox proportional hazard modelling, initially calculating the unadjusted hazard of each complication by age group, sex, index of multiple deprivation, ethnicity and modified Charlson co-morbidity index (Summary Hospital-level Mortality Indicator Specification; derived with maximum 5-year diagnosis code look-back period)^22–25^. Quintiles for index of multiple deprivation were derived from regional factors in England including average income, employment, education, housing, and crime. Quintile 1 includes the least deprived areas, quintile 5 includes the most deprived.

### Confounding factors

Cox proportional hazard models were adjusted for covariates which may have impacted the hazard of mortality. These included age, sex, ethnicity, index of multiple deprivation and Charlson co-morbidity index score. Unadjusted and adjusted hazard ratios were calculated (supplementary table 1).

### Sensitivity analysis

As ethnic group was missing in 9,358 procedures (14.3%) and no side was coded in 2,451 (3.7%) of procedures identified prior to the exclusion of non-diabetic amputations, these records were excluded (fig 1.) However, to investigate the impact of excluding these records the analysis was performed with these records included, and the primary outcomes and secondary outcomes were not significantly impacted by the exclusion of these records.

**Figure 1.**
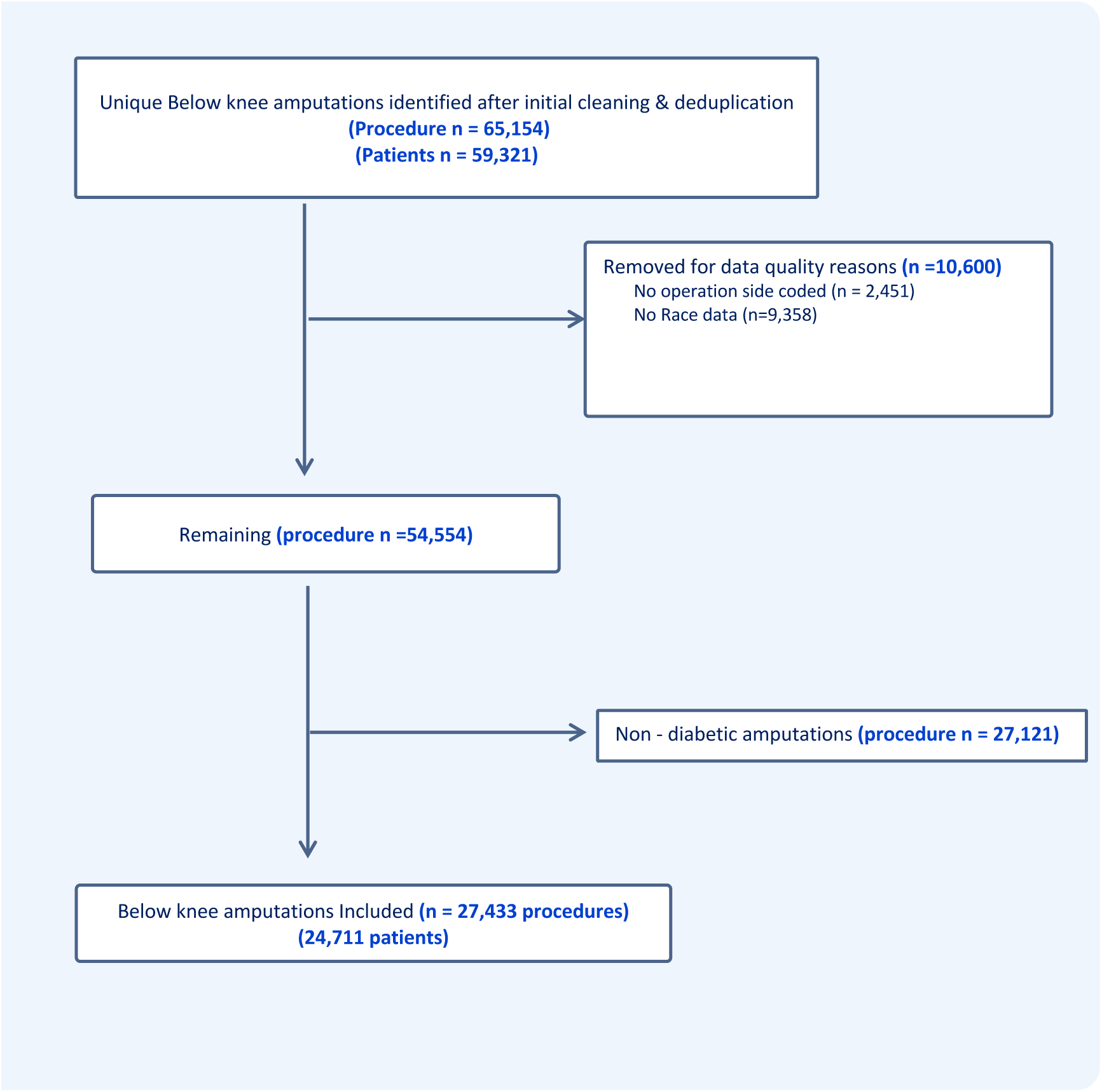
Data flowchart for hospital episode statistics (HES) data.

### Patient and public involvement

Four of the top 10 research priorities from the 2019 James Lind Alliance Priority Setting Partnership on Foot Health relate directly to neglected diabetic foot disease and its outcomes^26^. Patient representatives sit on the National Diabetes Foot Care Audit (NDFA) advisory group which includes clinicians, podiatrists, Diabetes UK and analysts from NHS England^6^. The advisory group provides advice and direction on future research with a recent emphasis on prevention of urgent amputation and the use of NHS databases. Our Diabetic MDT patient representatives reported that length of hospital stay and risks of surgery were important factors to consider in this study to improve shared decision making.

### Ethics

The nature of the data requested, the methods of analysis and dissemination, were discussed with NHS digital, and the University of Oxford Clinical Trials and Research governance team (CTRG). While the project was classified as research and has obtained university sponsorship under NDORMS, University of Oxford, it did not require ethical approval. No patient identifiable data is presented.

## Results

We identified 24,711 BKA on patients with diabetes in the 25-year period. The rate of BKA decreased from 2002 (5.9/100,000; 95% CI 5.6–6.1/100,000) to 2012 (4.4/100,000; 95% CI 4.2 – 4.5/100,000) and plateaued between 2012-2022 (4.3/100,000 in 2022; 95% CI 4.1 – 4.5/100,000). BKA rates were significantly higher in males (6.7/100,000; 95% CI 6.4 – 7/100,000) compared to females (2.0/100,000; 95% CI 1.8 – 2.1/100,000, P<0.05, Figure 2 and Table 1).

**Figure 2.**
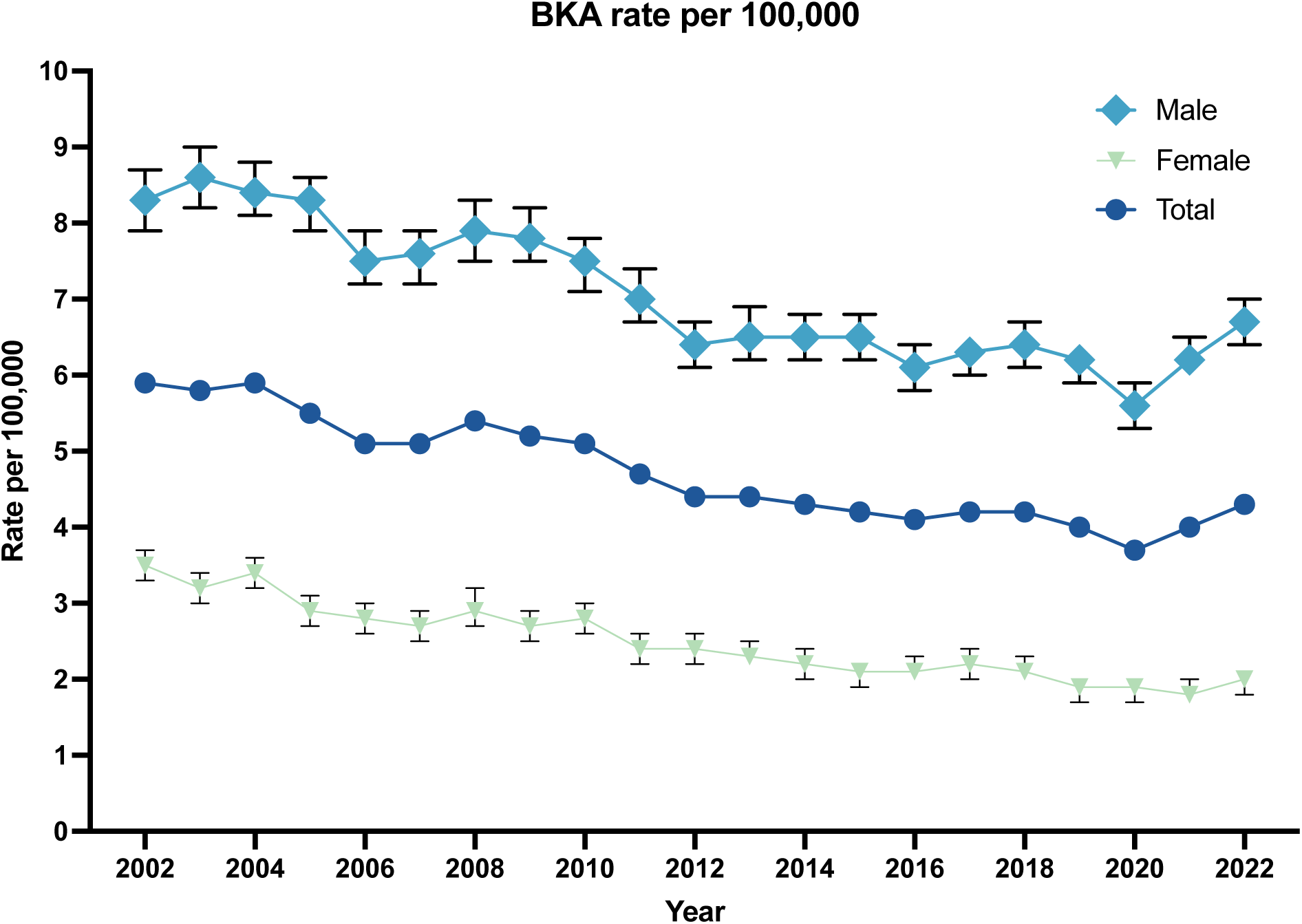
Rates of BKA per 100,00 of the population. Data shown is adjusted rate + 95% CI. The rate is age and sex standardized.

**Table 1.** Demographics and descriptive statistics of patient cohort.

|  | Number | % |
| --- | --- | --- |
| *Total Number* | 24,711 | 100% |
| *Sex* |  |  |
| *Male* | 18,835 | 76.22 |
| *Female* | 5,876 | 23.78 |
| *Race* |  |  |
| *White* | 23,275 | 94.19 |
| *Black* | 636 | 2.57 |
| *Asian* | 596 | 2.41 |
| *Mixed* | 54 | 0.22 |
| *Other* | 150 | 0.61 |
| *IMD* |  |  |
| *Most deprived 0-20%* | 8,397 | 33.98 |
| *More deprived 20-40%* | 5,683 | 23.00 |
| *Middle 40-60%* | 4,699 | 19.02 |
| *less deprived 60-80%* | 3,544 | 14.34 |
| *least deprived 80-100%* | 2,388 | 9.66 |
| *Age* |  |  |
| *0 - 19* | 7 | 0.03 |
| *20 - 39* | 818 | 3.31 |
| *40 - 59* | 7,386 | 29.89 |
| *60 - 79* | 13,651 | 58.24 |
| *80+* | 2,849 | 11.53 |
| *Charlson Group* |  |  |
| *Mild* | 754 | 3.18 |
| *Moderate* | 2,001 | 8.10 |
| *Severe* | 21,956 | 88.85 |

The mortality rates following BKA for diabetic foot disease were 11.3% at 90 days (2784 patients), 23.6% at 1 year (5817 patients), 58.2% at 5 years (in 8779 patients) and 92.3% at 25 years (Figure 3). A decrease in 90-day, 1-year and 2-year mortality rates were observed in England from 2002 to 2012 (Supplementary Figure 1). Since 2012 the corresponding 90-day, 1-year and 2-year mortality rates have not changed significantly.

**Figure 3.**
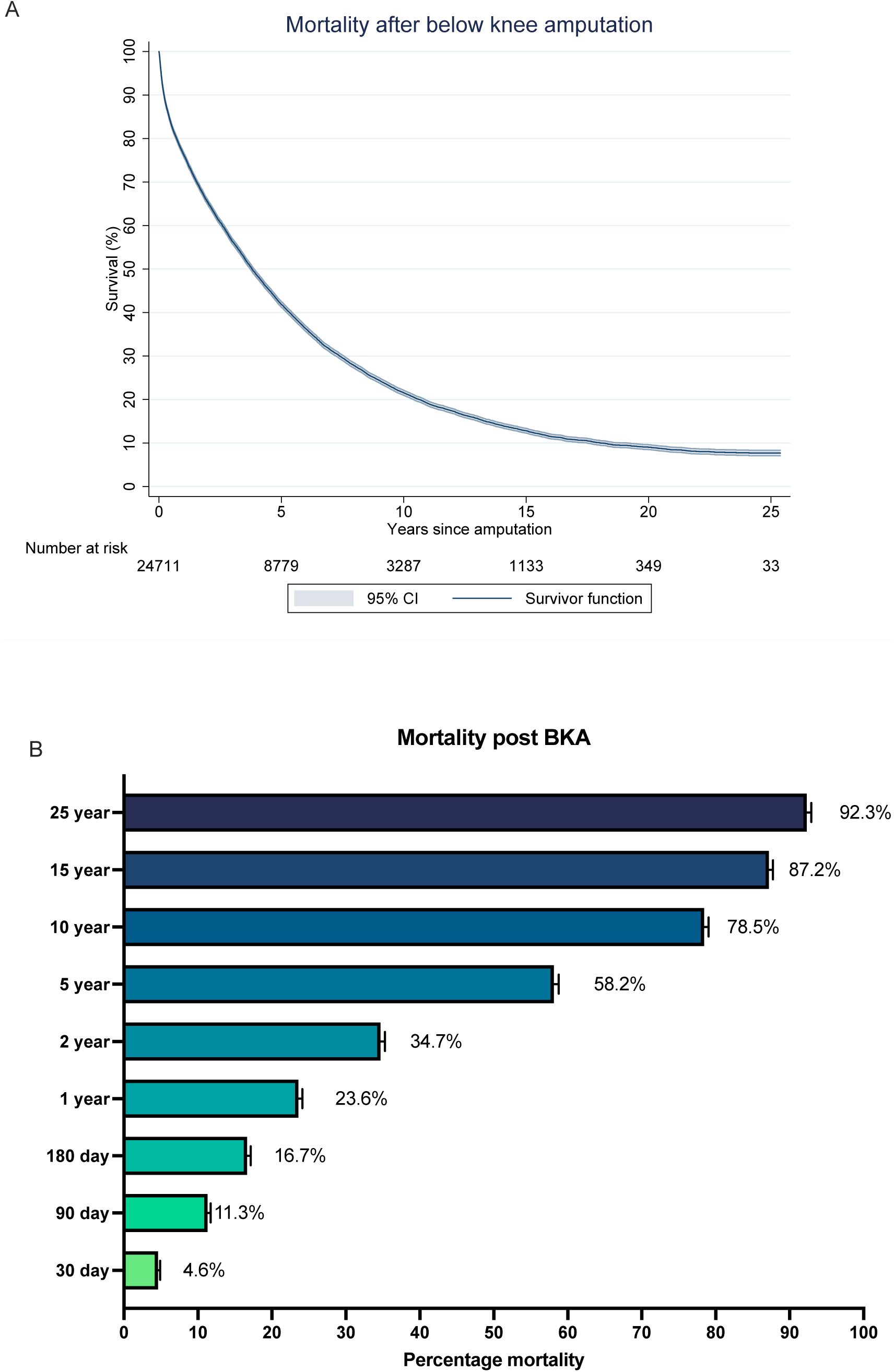
Survival analysis post below knee amputation in patients with diabetes. (A) Kaplan-Meier survival analysis. (B) Percentage mortality post below knee amputation.

The commonest causes of death were heart failure as a result of chromic ischemic heart disease (16.3%), acute myocardial infarction (10.0%), medical complications of poorly controlled diabetes (9.0%) and wound complications associated with the affected limb (7.7%, Figure 4). Cox proportional Hazard modelling (Figure 5) showed that female patients were at higher risk of mortality post BKA (HR 1.07 (95% CI 1.07 – 1.10). Increasing age was associated with higher mortality, with the 60-79 year old (HR 1.74 (95% CI 1.68 – 1.80)) and 80+ year old (HR 2.96 (95% CI 2.82 – 3.11) groups at higher risk of death post BKA. Increased Charlson co-morbidity index was associated with increased hazard of mortality, with ‘Moderate’ (HR 1.55 (1.04 – 2.32) and ‘Severe’ groups (HR 2.25 (95% CI 1.51 – 3.37) having significantly higher risk of mortality.

**Figure 4.**
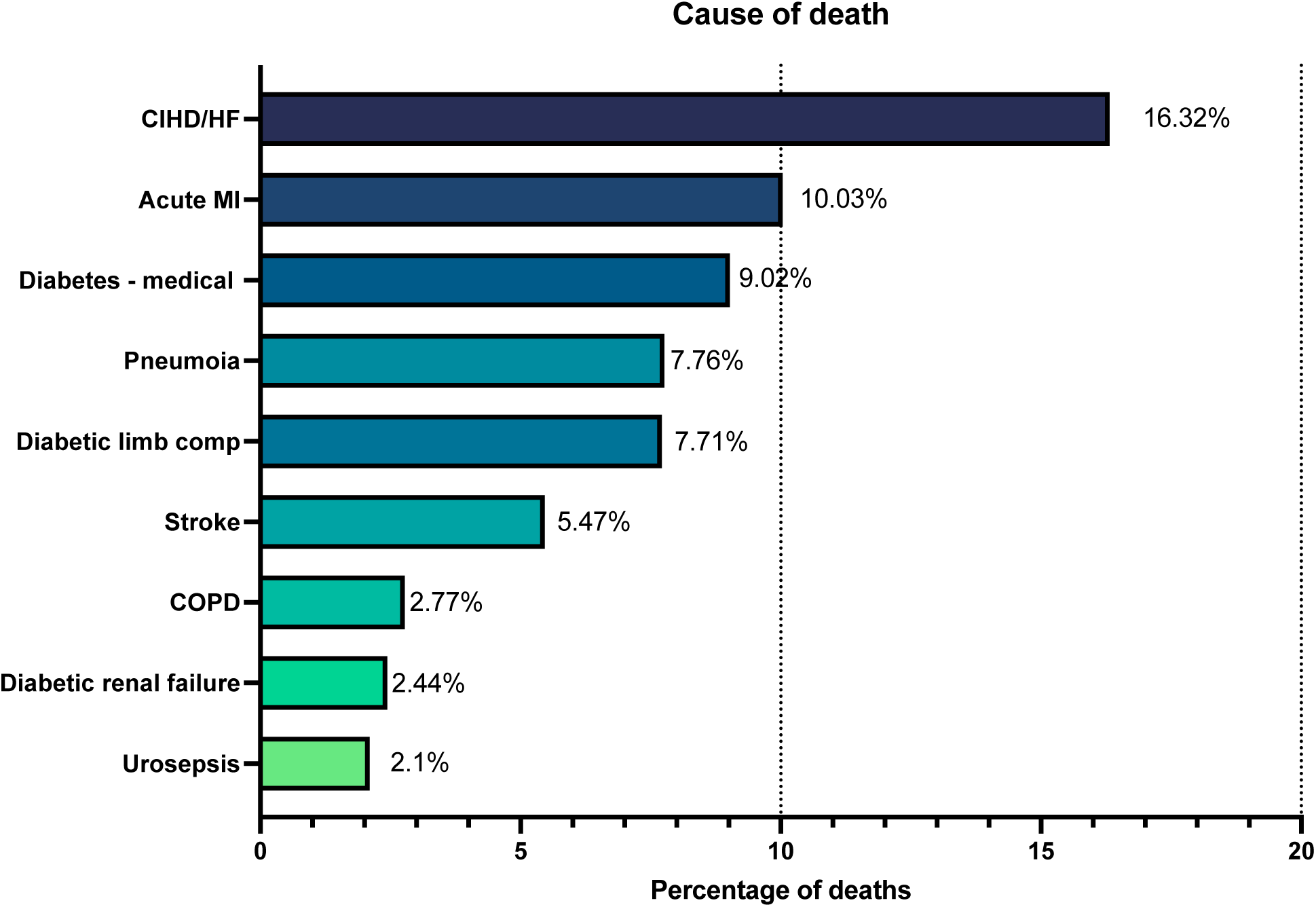
Cause of death in patients with diabetes post below knee amputation (1998-2023). Cause of death was acquired from ONS mortality data with the primary listed cause of death on the death certificate being listed.

**Figure 5.**
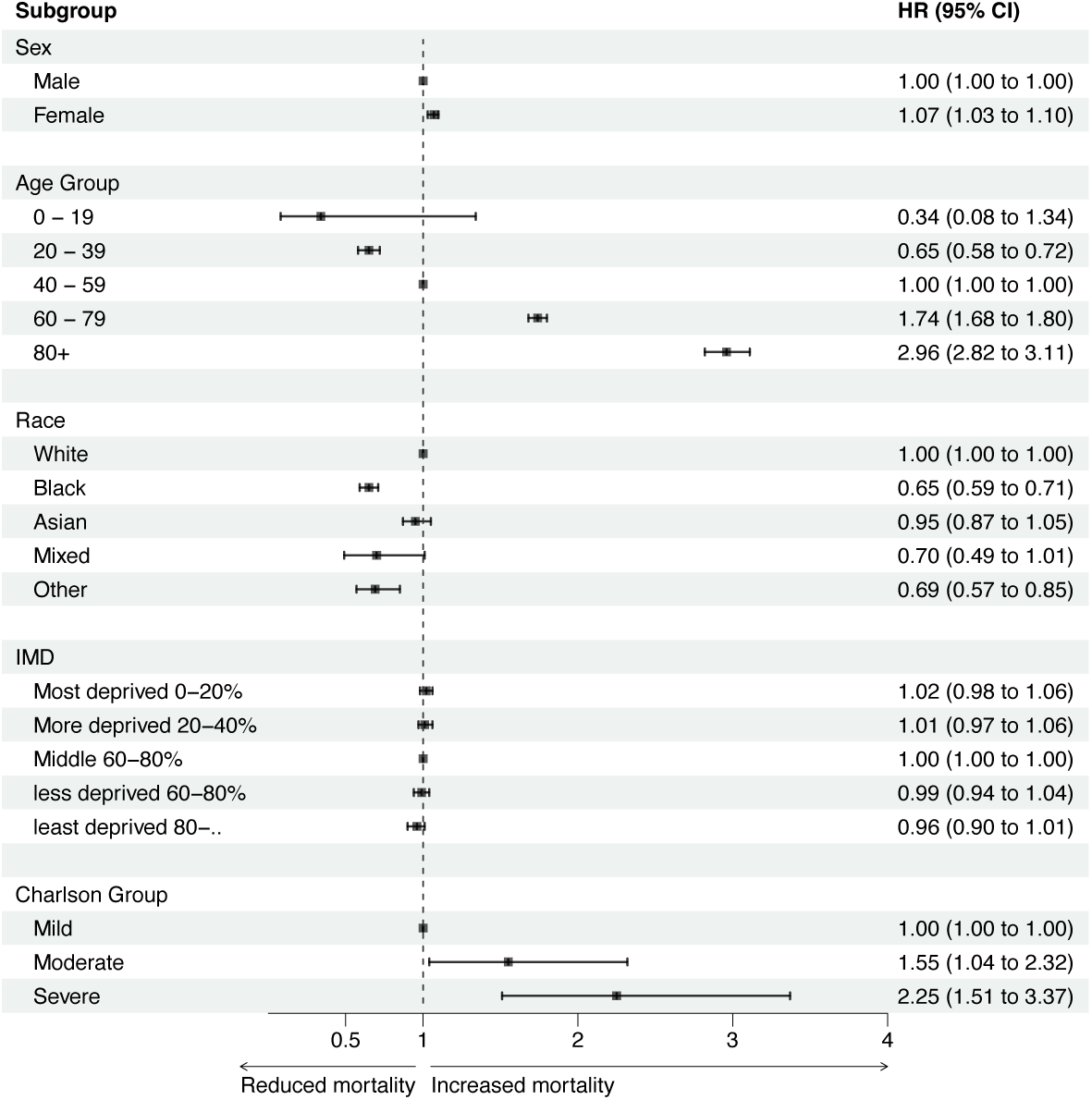
Hazard of mortality following below knee amputation. Forest plot summarizing cox regression analysis of demographic risk factors associated with mortality post below knee amputations. Data shown is adjusted HR, adjusted by sex, age group, Race/ethnicity, index of multiple deprivation (IMD) and Charlson comorbidity score group.

The 90-day reoperation rate for any cause was 10.4% (Table 2). Additional 90-day complications included acute kidney injury (12.1%), pulmonary embolus (0.59%, n=145), myocardial infarction (3.2%, n=799) and cerebrovascular accident (1.5%, n=367). The average length of stay was 33 days (IQR 29 days, range 1-152). 23.4% (n=5,717) of patients were readmitted to hospital within 90 days of discharge post BKA. Of these, 48.3% had a single readmission (n=2,762), 24.2% had two readmissions (n=1,381) and the remainder had three or more readmissions (27.5%, n=1574). Table 3 summarises the commonest causes of readmission.

**Table 2.** 90-day post operative complications, readmission rate and length of stay. LRTI – lower respiratory tract infection. UTI – urinary tract infection. MI – myocardial infarction. AKI – acute kidney injury. DVT – deep vein thrombosis. PE – Pulmonary embolism. Length of stay is calculated from day of admission in episode in which BKA is performed, until day of discharge.

|  | *n* | *%* | *95% CI* |
| --- | --- | --- | --- |
| **Total BKA numbers** | 24,711 |  |  |
| **Complications** |  |  |  |
| Any reoperation (ipsilateral leg) | 2,575 | 10.42 | (10.04 – 10.8) |
| 90-day readmission rate | 5717 | 23.14 | (22.6 – 23.6) |
| **90d Complications** |  |  |  |
| Mortality | 3269 | 11.32 | (10.92 – 11.71) |
| Reop. for Infection | 391 | 1.39 | (1.26 – 1.54) |
| AKI | 2989 | 12.09 | (11.7 – 12.5) |
| LRTI | 2778 | 11.24 | (10.85 – 11.6) |
| UTI | 2164 | 8.75 | (8.4 – 9.1) |
| MI | 799 | 3.23 | (3.02 – 3.46) |
| Stroke | 367 | 1.48 | (1.33 – 1.64) |
| DVT | 312 | 1.11 | (0.99 – 1.24) |
| PE | 145 | 0.59 | (0.49 – 0.69) |
| Fasciotomy | 90 | 0.32 | (0.25 – 0.39) |
|  | **(Mean)** | **Min-Max** | **IQR** |
| **Length of stay (days)** | 33 | 1-152 | 29 |

**Table 3.** Primary reason for admission if readmitted to hospital in first 90 days post discharge.

| ***90-day readmission reasons*** | ***% of readmissions*** |
| --- | --- |
| *Amputation wound complications* | 20.3 |
| *Uncontrolled diabetes* | 11.3 |
| *Sepsis* | 9.8 |
| *PVD* | 8.8 |
| *LRTI* | 6.4 |
| *Kidney* | 4.3 |
| *UTI* | 4.0 |
| *Heart failure* | 4.0 |
| *Ischemic heart disease* | 3.0 |
| *GI bleed* | 1.1 |
| *Cerebral vascular accident* | 0.9 |
| *Repeated falls* | 0.3 |
| *PE* | 0.3 |
| *Other* | 25.5 |

11.4% (95% CI 11. – 11.82%) underwent a contralateral amputation within 10 years of the index BKA (Figure 6).

**Figure 6.**
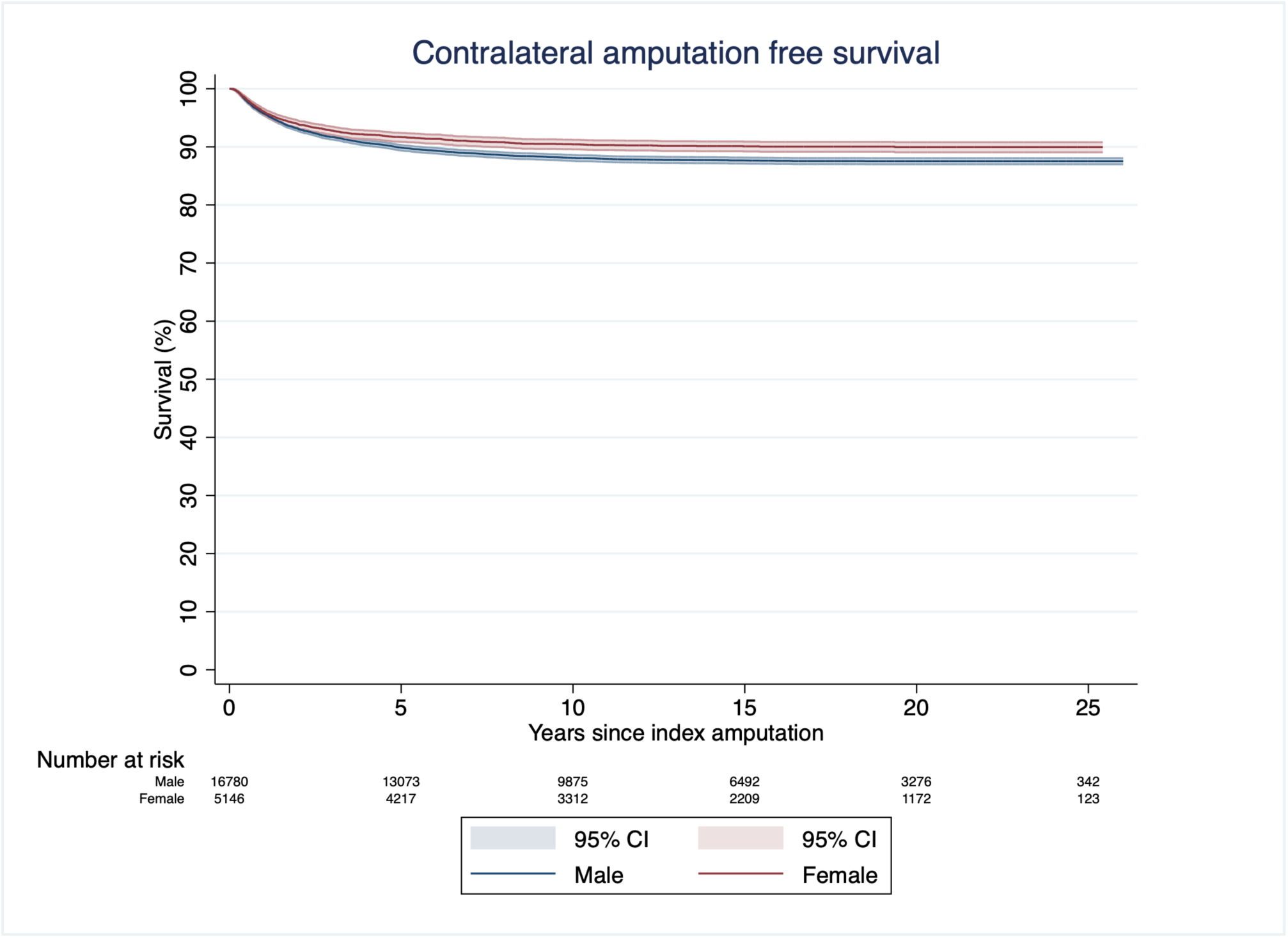
Kaplan Meier survival analysis of contralateral limb major amputation post below knee amputation.

## Discussion

The study demonstrates that the overall mortality associated with BKA in patients with diabetes in England is high and has not improved over the last 10 years. The 1-year mortality rate was 23.6%, increasing at 5 years to 58.2% and at 10 years to 78.4%. The operation itself is associated with a long in-patient hospital stay, a 90-day readmission rate of 24% and high rates of peri-operative medical and surgical complications. 10.4% underwent a more proximal ipsilateral re-amputation and 8.2% underwent a contralateral lower limb major amputation.

Our observations are comparable to those reported in the American Veterans cohort^9^ with similar 1-year mortality rate of 23%, rising to 72% in 7.5 years. This is despite significant differences between these two cohorts; the Veterans are almost entirely men, 40% did not have diabetes, and 27% are African-Americans. The Belgian Amputation Group most recently reported their national population based study of lower limb amputations from 2009 to 2018^27^. They defined ‘major’ lower limb amputations as any amputation proximal to the tarsus, comprising 13,247 of the 41,304 operations, of which 5,765 had diabetes. Kaplan-Meier survival analysis was limited to 5 years and revealed a mortality rate of 69% in patients with diabetes. This is higher than our observed mortality rate in England and may possibly be explained by the inclusion of AKA that are associated with higher mortality ^9 18^. A recent England population study of BKA and above knee amputation (AKA) for peripheral vascular disease also found that a subset of 13,943 BKA patients with diabetes had higher mortality rates than those PAD patients without diabetes^18^. The median survival post BKA of PAD patients with diabetes was 3.38 years compared to 5.72 years, and overall, AKA and BKA patients with diabetes had 20% greater mortality (95% CI 17% to 23%) than PAD patients without diabetes.

Together these large population studies emphasise the low survival probability of patients with diabetic foot disease who undergo BKA. Our findings suggest that the mortality rates have not improved over the last decade in patients with diabetes in England. We observe a decrease in 90-day, 1-year and 2-year mortality rates (but not the 5-year and 10-year mortality rates) until 2012, after which the rates plateau. A similar temporal trend is seen in 90-day mortality rate for PAD patients in England post AKA and BKA^18^. The shorter-term, in-hospital, mortality post lower limb amputation for PAD was found to decrease in England from 2003 to 2007^28^, in Germany from 2005 to 2009^29^, and in Spain from 2001 to 2012^30^. Interestingly, the Spanish study was the only one to analyse a subset of patients with diabetes and found no reduction in mortality. Similarly, one-year mortality rates remained stable for people with diabetes who underwent lower limb amputation between 2008 to 2014 in France^31^ and in Hong Kong (2001 - 2016) ^32^.

Strategies that reduce the high mortality include (i) reducing the incidence of BKA, (ii) preventing/treating the common causes of death, and/or (iii) identifying at risk individuals to re- direct healthcare resources.

There has been a UK drive to reduce the incidence of BKA by improving the care of diabetic foot ulcers, including the creation of Multi-disciplinary Foot Care Services (MFCS) following NICE guidelines (NG19) in 2015 ^7^. Our analysis of HES data from the preceding decade saw a much greater decline in BKA rate, particularly in men, until 2012 and thereafter the rates have remained at approximately 4.5/100,000. The most recent UK National Diabetes Foot Care Audit reported improvements in ulcer healing rates and an increase in recorded diabetic foot ulcers ‘episodes’ from 2014 to 2021^6^. What is not clear is whether the emphasis on earlier ulcer treatment with more minor surgery has an impact on BKA rates and survival. A limitation of this study is that it does not examine the number and type of previous ipsilateral lower limb operations that were performed.

Improvements in the incidence of BKA may have been offset by the increased incidence of diabetes in general, and the subsequent development of ulcers in particular. There are signs that the NHS is now struggling to meet the growing demand. The latest data from the National Diabetes Foot Audit (NDA) showed that 66.3% of people with diabetic foot ulcer were seen by a foot specialist within the recommended 13 days during 2022/23. For the first time, this represents a decrease from the previous year’s 69%^6^.

Mortality post BKA for diabetes could be improved by identifying individuals most at risk of death. Our analysis revealed that female sex, increasing age, and higher co-morbidity indices were associated with a significantly higher mortality rate. Cardio- and peripheral vascular disease, kidney disease, sepsis and uncontrolled diabetes were the commonest causes of both death and readmission to hospital (Figure 3, Table 3). Nearly one in four patients were readmitted within 90 days post index BKA at great personal and societal cost. The average length of stay was over a month. We would argue that this long post-operative period provides a critical opportunity for urgent in-patient healthcare intervention to optimise blood sugar control, cardiovascular risk factors and peripheral vascular disease. There is precedence for medical intervention post urgent surgery in at risk group, for example the mortality associated with hip fractures has been successfully reduced by a nationwide change to ortho-geratology led MDT care^33^. Our findings suggest that the peri-operative period of BKA may be the last opportunity to intervene in this highly vulnerable group.

In our data, BKA was associated with worst outcome in those patients with the highest co-morbidity index, irrespective of race. The study is limited to a national analysis and further work will investigate if ethnic groups in specific areas are particularly disadvantaged.

The observations of this study are further limited by the accuracy of the recorded OPCS-4 and ICD-10 codes. The results and conclusions drawn are susceptible to misclassification or omission of codes. Our study does not distinguish between type 1 or type 2 diabetes. It also does not distinguish between different reasons for BKA in the diabetic population. All patients with diabetes that underwent a BKA were included in this study, but that does not mean that all patients underwent BKA for a diabetic foot infection. We excluded patients with OPCS-4 or ICD-10 codes pertaining to trauma, tumour and/or sarcoma, but there remains the possibility that a minority of patients underwent BKA for non diabetic foot infection reasons e.g. complex regional pain syndrome.

## Conclusions

This landmark 25-year England population study has revealed that BKA was associated with high mortality rates showing little improvement over the last 10 years. Severely co-morbid individuals, females and people older than 60 years had the highest mortality rates and represent at risk groups. BKA was associated with high peri-operative morbidity, high rate of further amputation and high incidence of significant complications. Nearly one in four patients were readmitted within the first 90 days. Collectively these observations depict a severely comorbid and vulnerable population whose surgical admission may be one of the last opportunities for urgent healthcare intervention.

## Data Availability

All data produced in the present study are available upon reasonable request to the authors

---

**Supplementary Figure 1.**
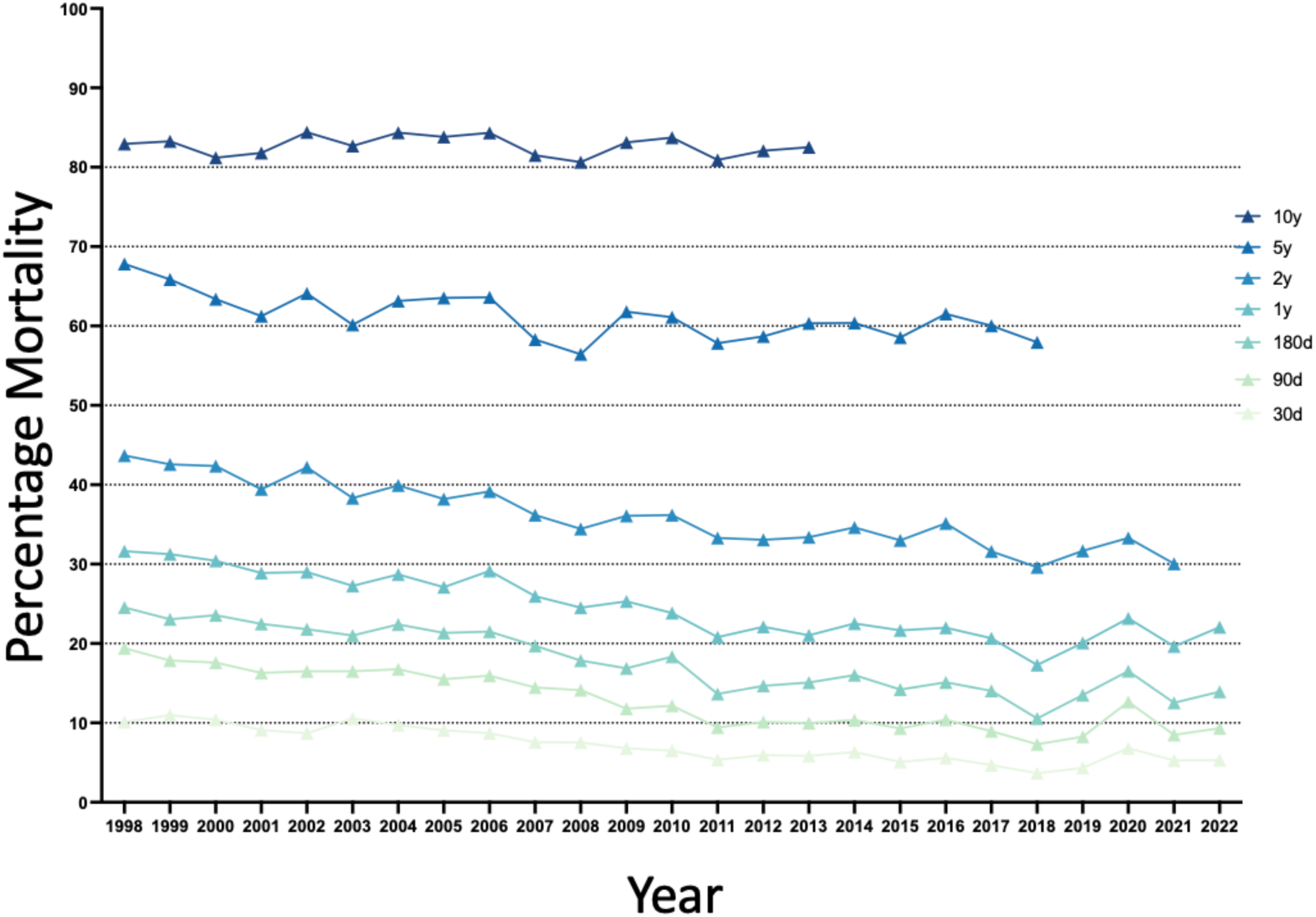
Mortality post BKA in England over time. 30-day, 90-day, 180-day, 1-year, 2-year, 5-year and 10-year mortality post BKA over 25 years in England.

**Supplementary table 1.** Unadjusted and adjusted hazard ratios of mortality following below knee amputation.

|  | *Unadjusted*<br>*HR* | *95% CI* | *Adjusted*<br>*HR* | *95% CI* |
| --- | --- | --- | --- | --- |
| **Sex** |  |  |  |  |
| Male | 1 |  |  |  |
| Female | 1.084347 | (1.05 - 1.12) | 1.067838 | (1.03 - 1.10) |
| **Age group** |  |  |  |  |
| 0 - 19 | 0.2848151 | (0.07 - 1.14) | 0.3351812 | (0.08 - 1.34) |
| 20 - 39 | 0.6374038 | (0.58 - 0.70) | 0.6466232 | (0.58 - 0.72) |
| 40 - 59 | 1 |  |  |  |
| 60 - 79 | 1.794464 | (1.73 - 1.86) | 1.742359 | (1.68 - 1.80) |
| 80+ | 3.129881 | (2.98 - 3.29) | 2.959687 | (2.82 - 3.11) |
| **Race** |  |  |  |  |
| White | 1 |  | 1 |  |
| Black | 0.7635454 | (0.69 - 0.84) | 0.6481803 | (0.59 - 0.71) |
| Asian | 0.9899374 | (0.90 - 1.09) | 0.9548381 | (0.87 - 1.05) |
| Mixed | 0.7092968 | (0.49 - 1.02) | 0.7040646 | (0.49 - 1.01) |
| Other | 0.8078466 | (0.66 - 0.99) | 0.6949492 | (0.57 - 0.85) |
| **IMD** |  |  |  |  |
| Most deprived 0-20% | 0.9051064 | (0.87 - 0.94) | 1.018346 | (0.98 - 1.06) |
| More deprived 20-40% | 0.9650289 | (0.92 - 1.01) | 1.013097 | (0.97 - 1.06) |
| Middle 60-80% | 1 |  | 1 |  |
| less deprived 60-80% | 1.026245 | (0.98 - 1.08) | 0.9923189 | (0.94 - 1.04) |
| least deprived 80-100% | 1.02534 | (0.97 - 1.08) | 0.9564321 | (0.90 - 1.01) |
| **Charlson Group** |  |  |  |  |
| Mild | 1 |  | 1 |  |
| Moderate | 1.751544 | (1.17 - 2.62) | 1.553416 | (1.04 - 2.32) |
| Severe | 2.599769 | (1.74 - 3.88) | 2.253103 | (1.51 - 3.37) |

## References

1. Walters DP, Gatling W, Mullee MA, et al. The distribution and severity of diabetic foot disease: a community study with comparison to a non-diabetic group. Diabet Med 1992;9(4):354–8. doi: 10.1111/j.1464-5491.1992.tb01796.x

2. Ramsey SD, Newton K, Blough D, et al. Incidence, outcomes, and cost of foot ulcers in patients with diabetes. Diabetes Care 1999;22(3):382–7. doi: 10.2337/diacare.22.3.382

3. Siitonen OI, Niskanen LK, Laakso M, et al. Lower-Extremity Amputations in Diabetic and Nondiabetic Patients: A population-based study in eastern Finland. Diabetes Care 1993;16(1):16–20. doi: 10.2337/diacare.16.1.16

4. Van Damme H, Limet R. Amputation in Diabetic Patients. Clinics in Podiatric Medicine and Surgery 2007;24(3):569–82. doi: 10.1016/j.cpm.2007.03.007

5. Ahmad N, Thomas GN, Gill P, et al. The prevalence of major lower limb amputation in the diabetic and non-diabetic population of England 2003-2013. Diabetes & Vascular Disease Research 2016;13(5):348–53. doi: 10.1177/1479164116651390

6. Audit NDFC. National Diabetes Foot Care Audit https://digitalnhsuk/data-and-information/clinical-audits-and-registries/national-diabetes-foot-care-audit 2023

7. 19 NG. Diabetic foot problems: prevention and management. https://wwwniceorguk/guidance/ng19 2015

8. Mayfield JA, Reiber GE, Maynard C, et al. Survival following lower-limb amputation in a veteran population. Journal of Rehabilitation Research and Development 2001;38(3):341–45.

9. Feinglass J, Pearce WH, Martin GJ, et al. Postoperative and late survival outcomes after major amputation: Findings from the Department of Veterans Affairs National Surgical Quality Improvement Program. Surgery 2001;130(1):21–29. doi: 10.1067/msy.2001.115359

10. Cruz CP, Eidt JF, Capps C, et al. Major lower extremity amputations at a Veterans Affairs hospital. American Journal of Surgery 2003;186(5):449–54. doi: 10.1016/j.amjsurg.2003.07.027

11. Bhatnagar V, Richard E, Melcer T, et al. Retrospective study of cardiovascular disease risk factors among a cohort of combat veterans with lower limb amputation. Vascular Health and Risk Management 2019;15:409–18. doi: 10.2147/vhrm.S212729

12. Hennessy C, Pring T, Qureshi B, et al. 1141 Mortality and Re-Amputation Rates Following Below Knee Amputation: A Systematic Review and Meta-Analysis. British Journal of Surgery 2024;111(Supplement_6) doi: 10.1093/bjs/znae163.005

13. Lavery LA, Hunt NA, Ndip A, et al. Impact of Chronic Kidney Disease on Survival After Amputation in Individuals With Diabetes. Diabetes Care 2010;33(11):2365–69. doi: 10.2337/dc10-1213

14. Singh RK, Prasad G. Long-term mortality after lower-limb amputation. Prosthet Orthot Int 2016;40(5):545–51. doi: 10.1177/0309364615596067 [published Online First: 20150807]

15. Fortington LV, Geertzen JHB, van Netten JJ, et al. Short and Long Term Mortality Rates after a Lower Limb Amputation. European Journal of Vascular and Endovascular Surgery 2013;46(1):124–31. doi: 10.1016/j.ejvs.2013.03.024

16. Karam J, Shepard A, Rubinfeld I. Predictors of operative mortality following major lower extremity amputations using the National Surgical Quality Improvement Program public use data. Journal of Vascular Surgery 2013;58(5):1276–82. doi: 10.1016/j.jvs.2013.05.026

17. Ahmad N, Thomas GN, Gill P, et al. Lower limb amputation in England: prevalence, regional variation and relationship with revascularisation, deprivation and risk factors. A retrospective review of hospital data. Journal of the Royal Society of Medicine 2014;107(12):483–89. doi: 10.1177/0141076814557301

18. Maheswaran R, Tong T, Michaels J, et al. Time trends and geographical variation in major lower limb amputation related to peripheral arterial disease in England. BJS Open 2024;8(1) doi: 10.1093/bjsopen/zrad140

19. Statistics. NDHE. https://digitalnhsuk/data-and-information/data-tools-and-services/data-services/hospital-episode-statistics

20. Quan H, Li B, Couris CM, et al. Updating and Validating the Charlson Comorbidity Index and Score for Risk Adjustment in Hospital Discharge Abstracts Using Data From 6 Countries. American Journal of Epidemiology 2011;173(6):676–82. doi: 10.1093/aje/kwq433

21. Herbert A, Wijlaars L, Zylbersztejn A, et al. Data Resource Profile: Hospital Episode Statistics Admitted Patient Care (HES APC). Int J Epidemiol 2017;46(4):1093–93i. doi: 10.1093/ije/dyx015

22. Noble MW, G · Smith, G ·, et al. Measuring multiple deprivation at the small-area level. Environ Plan A 2006;38(169-185)

23. Digital N. Summary hospital-level mortality indicator (SHMI). Version 1.25. https://wwwdigitalnhsuk/SHMI July 2017

24. Zhang JX, Iwashyna TJ, Christakis NA. The performance of different lookback periods and sources of information for Charlson comorbidity adjustment in Medicare claims. Med Care 1999;37(11):1128–39. doi: 10.1097/00005650-199911000-00005

25. Charlson ME, Pompei P, Ales KL, et al. A new method of classifying prognostic comorbidity in longitudinal studies: development and validation. J Chronic Dis 1987;40(5):373–83. doi: 10.1016/0021-9681(87)90171-8

26. PSP JLA. Foot Health Priorities. https://wwwjlanihracuk/priority-setting-partnerships/foot-health#tab-31511 2019

27. Lauwers P, Wouters K, Vanoverloop J, et al. The impact of diabetes on mortality rates after lower extremity amputation. Diabetic Medicine 2024;41(1) doi: 10.1111/dme.15152

28. Moxey PW, Hofman D, Hinchliffe RJ, et al. Epidemiological study of lower limb amputation in England between 2003 and 2008. British Journal of Surgery 2010;97(9):1348–53. doi: 10.1002/bjs.7092

29. Malyar NM, Freisinger E, Meyborg M, et al. Low Rates of Revascularization and High In-Hospital Mortality in Patients With Ischemic Lower Limb Amputation: Morbidity and Mortality of Ischemic Amputation. Angiology 2016;67(9):860–69. doi: 10.1177/0003319715626849

30. Lopez-de-Andres A, Jiménez-García R, Aragón-Sánchez J, et al. National trends in incidence and outcomes in lower extremity amputations in people with and without diabetes in Spain, 2001–2012. Diabetes Research and Clinical Practice 2015;108(3):499–507. doi: 10.1016/j.diabres.2015.01.010

31. Amadou C, Denis P, Cosker K, et al. Less amputations for diabetic foot ulcer from 2008 to 2014, hospital management improved but substantial progress is still possible: A French nationwide study. PLoS One 2020;15(11):e0242524. doi: 10.1371/journal.pone.0242524 [published Online First: 20201130]

32. Wu H, Yang A, Lau ESH, et al. Secular trends in rates of hospitalisation for lower extremity amputation and 1 year mortality in people with diabetes in Hong Kong, 2001–2016: a retrospective cohort study. Diabetologia 2020;63(12):2689–98. doi: 10.1007/s00125-020-05278-2

33. Van Heghe A, Mordant G, Dupont J, et al. Effects of Orthogeriatric Care Models on Outcomes of Hip Fracture Patients: A Systematic Review and Meta-Analysis. Calcif Tissue Int 2022;110(2):162–84. doi: 10.1007/s00223-021-00913-5 [published Online First: 20210930]

